# COVID-associated pediatric hospitalization and ICU admission trends across a multi-state health system and the broader US population

**DOI:** 10.1101/2021.04.02.21254593

**Authors:** AJ Venkatakrishnan, Colin Pawlowski, John C. O’Horo, Andrew Badley, John Halamka, Venky Soundararajan

**Author notes:** Correspondence to Venky Soundararajan. Joint first authors (AJ Venkatakrishnan, Colin Pawlowski).

## Abstract

Public health concerns are emerging based on reports of new SARS-CoV-2 variant strains purportedly triggering a rise in COVID-associated hospitalizations and ICU admissions, particularly in younger patients and the pediatric population. However, analyzing health records of COVID patients from the electronic health records (EHRs) of a multi-state US healthcare system, we find that there is actually a significant drop in COVID-associated hospitalization rates and ICU admission rates in March 2021 compared to February 2021. We further triangulate these EHR-derived insights with the official US government epidemiological data sets to show that during this same time period, there is no apparent nation-wide spike in pediatric hospitalizations. Our study motivates the need to develop a real-time system that integrates various COVID hospitalization and ICU monitoring efforts from the EHR databases of various health systems together with national epidemiological data sets. By infusing SARS-CoV-2 genomic sequencing data to flag potentially new or emergent viral strains, as well as county-level COVID vaccine rollout rates and shifts in SARS-CoV-2 PCR positivity rates into such a real-time monitoring system, public health policies and media reporting can be more effectively informed through the rigor of holistic biomedical data sciences.

## Introduction

Media reports this week are suggesting a potential spike up in COVID-19 genetic variants-associated intensive care unit (ICU) admissions for younger patients from the continents of North America^1–3^, South America^4,5^, and Europe^6,7^. Here, we conducted an observational study on the health records of 60,539 patients (March 1-28, 2021) and 60,256 patients (February 1-28, 2021) from the multi-state Mayo Clinic health system for whom the SARS-CoV-2 PCR test results were also available. We then juxtaposed these results on to the national epidemiological data from the US government as of March 29, 2021 (https://healthdata.gov/).^8^

## Methods

### Study design

This is a retrospective study of individuals who underwent polymerase chain reaction (PCR) testing for suspected SARS-CoV-2 infection at the Mayo Clinic and hospitals affiliated with the Mayo Clinic Health System. This study was reviewed by the Mayo Clinic Institutional Review Board (IRB) and determined to be exempt from the requirement for IRB approval (45 CFR 46.104d, category 4). Subjects were excluded if they did not have a research authorization on file.

## Results

Analysis of the health records from Mayo Clinic health system for patients where the SARS-CoV-2 PCR test results were also available as of March 29, 2021 shows a significant drop in the ICU admission rates during March (1.45%; 87 of 6017 COVID_pos_ patients) compared to February (1.99%; 121 of 6078 COVID_pos_ patients) with a Rate Ratio of 0.78 (p-value = 0.0002; **Figure 1a**). In this study population, there is a slight shift in the peak of ICU admissions towards the younger population in March relative to February (**Figure 1b**). While this trend is not statistically significant at this time, this particular shift requires further monitoring over the coming weeks.

**Figure 1.**
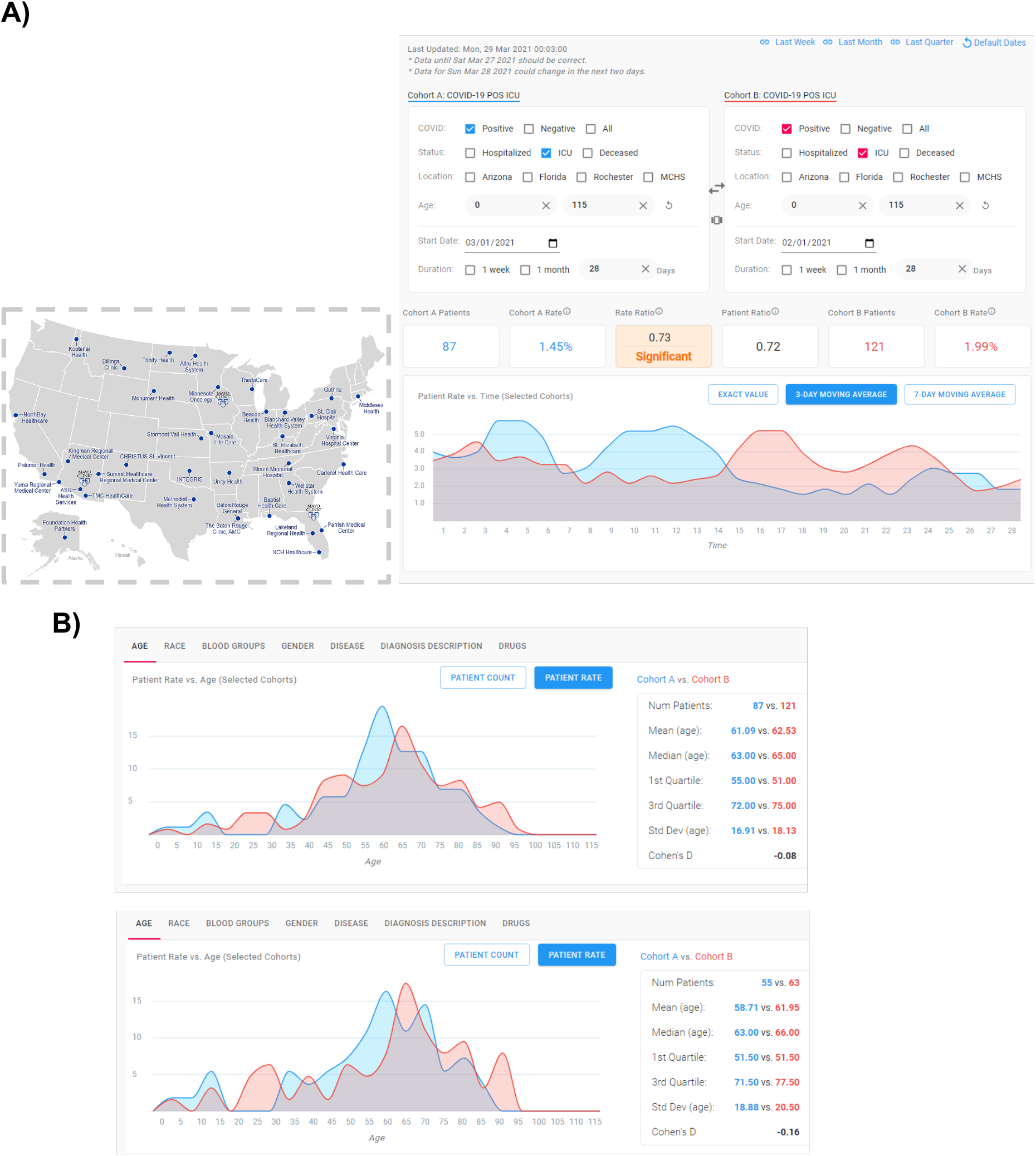
**(A)** Summary of patient electronic health records across the Mayo Clinic health system sites (*boxed insert*) for patients diagnosed with COVID-19 shows a significantly lower level of ICU admissions in the first four weeks of March 2021 (n = 87 ICU patients from March 1 to March 28; *blue*) compared to the comparable four week period of February 2021 (n = 121 ICU patients from February 1 to February 28; *red*). This significant decline in COVID-induced ICU admissions in March relative to the comparable period of February 2021 has an associated Rate Ratio of 0.73. The ICU admission rate is computed as a proportion of positive COVID diagnosis. **(B)** Slight shift in the peak towards younger patients seen in ICU admissions for COVID-positive (COVID_pos_) patients in March (blue) relative to February (red). *Top panel -* Data from all sites constituting Mayo Clinic health system is shown here. *Bottom panel -* Same analysis but restricted to the Rochester MN and Mayo Clinic health system sites (without considering the Arizona and Florida sites). This minor shift to younger populations requires further monitoring over the coming weeks.

Restricting the study population to younger COVID_pos_ patients (age < 55), we observe a drop in the number of ICU admissions in March compared to February (not statistically significant; **Figure 2a**). Consistent with the drop in the ICU admission rate, we also see a statistically significant drop in the hospitalization rate in March 2021 (10.97%; 660 of 6017 COVID_pos_ patients) compared to February 2021 (13.64%; 829 of 6078 COVID_pos_ patients) with a rate ratio of 0.8 (p-value: 2.34e-12; **Figure 2b**). Focusing the study population to the younger COVID_pos_ patients (age < 55), we observe a drop in the number of hospitalizations in March compared to February (not statistically significant; **Figure 2c**).

**Figure 2.**
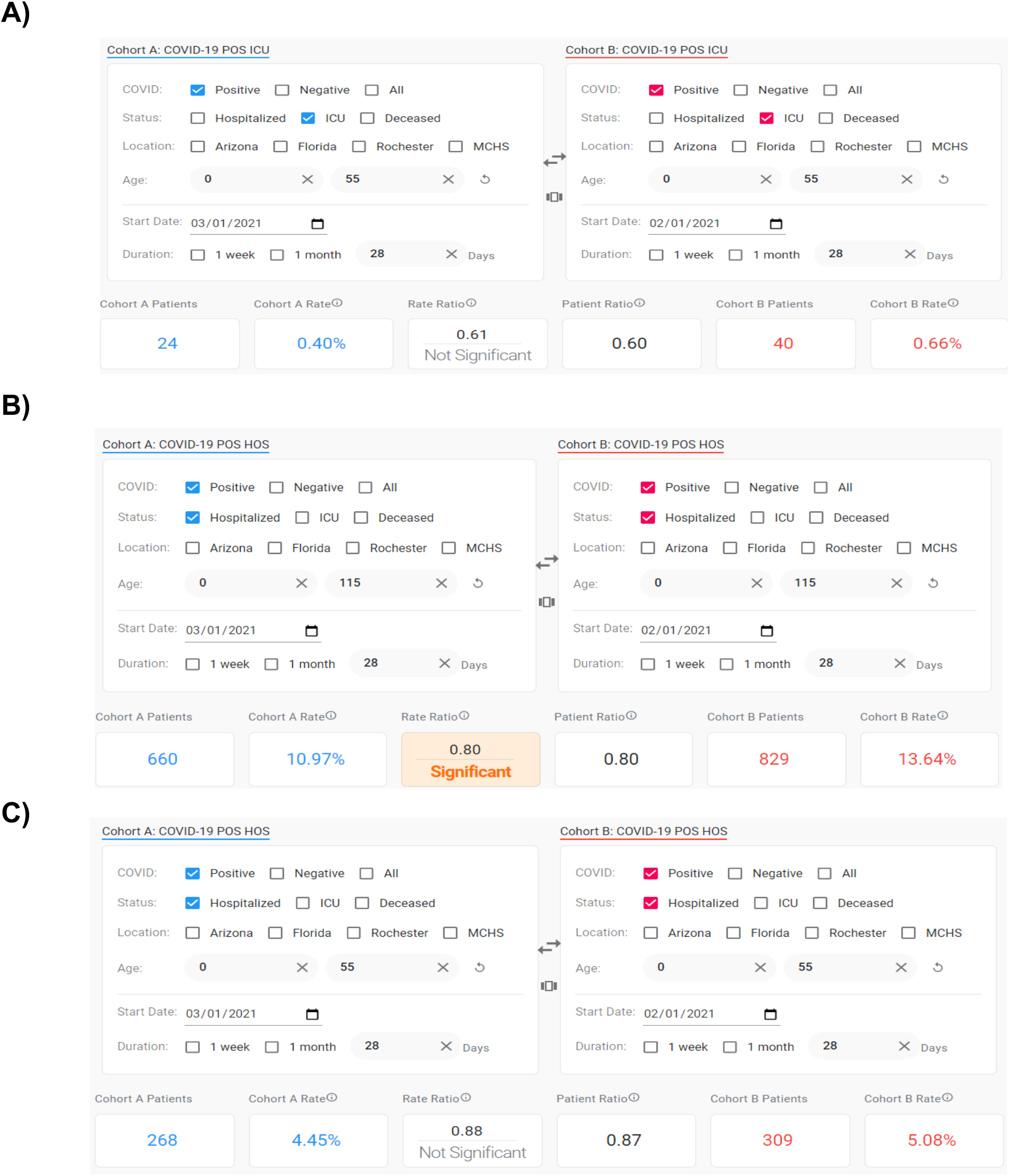
**(A)** Comparing March and February ICU admissions for younger (age <55) COVID_pos_ patients at the Mayo Clinic and associated health system shows a drop in the ICU admission rate in March relative to February 2021 (not statistically significant). The ICU admission rate is computed as a proportion of positive COVID diagnosis. **(B)** Comparing March and February hospitalization rate for COVID-positive (COVID_pos_) patients at the Mayo Clinic and associated health system shows a statistically significant drop in March compared to February. The hospitalization rate is computed as a proportion of positive COVID diagnosis. (**C)** Comparing March and February hospitalization for younger (age <55) COVID_pos_ patients at the Mayo Clinic and associated health system shows a drop in the hospitalization rate in March relative to February 2021 (not statistically significant). The ICU rate is computed as a proportion of positive COVID diagnosis.

Independently, the national data from the US government (https://healthdata.gov/)^8^ also shows a steady drop over recent months in hospitalized pediatric patients with confirmed or suspected COVID-19 (**Figure 3a**). We observe similar trends in the top-10 states by population as well (**Figure 3b**).

**Figure 3.**
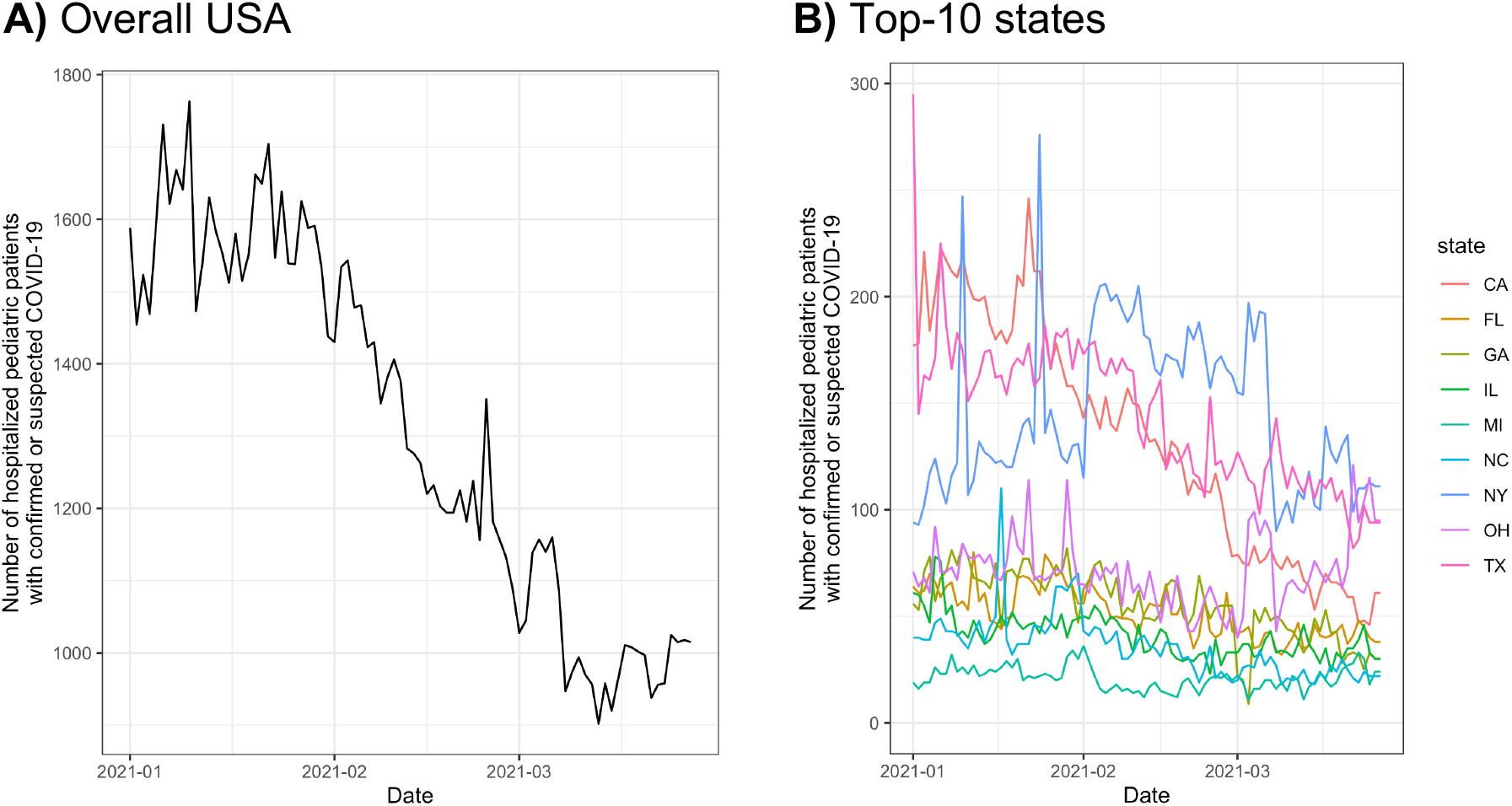
**(A)** National and state-level counts of hospitalized pediatric patients with confirmed or suspected COVID-19 from January 1, 2021 to March 27, 2021 (Overall trend for USA). **(B)** Trends for the top-10 most populous states, including: California, Florida, Georgia, Illinois, Michigan, North Carolina, New York, Ohio, and Texas.

## Discussion

This preliminary analysis shows that there is a decreasing trend in ICU admissions and hospitalization of COVID-19 patients across a large multi-state health system as of March 2021. Furthermore, there is no apparent nation-wide spike in pediatric hospitalizations in March compared to February 2021. Decreased hospitalizations and ICU admission rates may be reflective of advances in early diagnosis and intervention. As the proportion of circulating viruses shift towards more variants, these results may change. Overall, amidst recent concerns about emergent SARS-CoV-2 strains, there is an imminent need to develop a real-time system that integrates various national surveillance efforts in order to guide public health policies.

## Data Availability

After publication, the data will be made available to others upon reasonable requests to the corresponding author. A proposal with a detailed description of study objectives and the statistical analysis plan will be needed for evaluation of the reasonability of requests.

